# Distinguishing recrudescence from reinfection in lymphatic filariasis: a genomics-based approach for monitoring worm burden

**DOI:** 10.1101/2023.11.05.23297155

**Authors:** Young-Jun Choi, Kerstin Fischer, Aboulaye Méité, Benjamin G. Koudou, Peter U. Fischer, Makedonka Mitreva

**Author notes:** Correspondence: Makedonka Mitreva. Emails: Young-Jun Choi, Kerstin Fischer, Aboulaye Méité, Benjamin G. Koudou, Peter U. Fischer, Makedonka Mitreva.

## Abstract

**Background:** The Global Program to Eliminate Lymphatic Filariasis is the largest public health program based on mass drug administration (MDA). Despite decades of MDA, ongoing transmission in some countries remains a challenge. To optimize interventions, it is essential to differentiate between recrudescence (poor drug response and persistent infection) and new infections (ongoing transmission). Since adult filariae are inaccessible in humans, an approach that relies on genotyping the offspring microfilariae (mf) is required.

**Methods:** We utilized *Brugia malayi* adults and mf obtained from gerbils with a known pedigree to develop and validate our whole-genome amplification and kinship analysis approach. We then sequenced the genomes of *Wuchereria bancrofti* mf from infected humans from Côte d’Ivoire (CDI), characterized the population genetic diversity, and made inferences about the adult breeders. We developed a whole-exome capture panel for *W. bancrofti* to enrich parasite nuclear DNA from lower-quality samples contaminated with host DNA.

**Results:** We established a robust analysis pipeline using *B. malayi* adult and mf. We estimated the pre-treatment genetic diversity in *W. bancrofti* from 269 mf collected from 18 individuals, and further analyzed 1-year post-treatment samples of 74 mf from 4 individuals. By reconstructing and temporally tracking sibling relationships across pre- and post-treatment samples, we differentiated between new and established maternal families, suggesting reinfection in one subject and recrudescence in three subjects. Estimated reproductively active adult females ranged between 3 and 9 in the studied subjects. Hemizygosity of the male X-chromosome allowed for direct inference of haplotypes, facilitating robust maternal parentage inference, even when the genetic diversity was low. Population structure analysis revealed genetically distinct parasites among our CDI samples. Sequence composition and variant analysis of whole-exome libraries showed that the hybridization capture approach can effectively enrich parasite nuclear DNA and identify protein-coding variants with ∼95% genotype concordance rate.

**Conclusions:** We have generated resources to facilitate development of field-deployable genotyping tools that can estimate worm burdens and monitor parasite populations. These tools are essential for the success of lymphatic filariasis MDA programs. With further expansion of the databases to include geographically diverse samples, we will be able to spatially track parasite movement associated with host/vector migration.

## Background

Population genomic approaches are continually transforming the field of infectious disease by enabling rigorous examination of epidemiological processes such as pathogen migration, strain divergence, and selection [1, 2]. For parasitic infections, genomic surveillance through spatiotemporal analysis of the distribution of genetic variation within and between hosts and geographic regions can uncover transmission dynamics and the impact of intervention strategies, such as mass drug administration (MDA), on parasite populations [3]. Lymphatic filariasis (LF) is a widespread mosquito-borne parasitic disease targeted for global elimination by the World Health Organization [4], and has remained relatively untouched by the advances in genomic approaches compared to other pathogen groups, despite its global health significance [4, 5]. One reason that progress has been difficult is that the adult stages of the LF-causing nematodes (*Wuchereria bancrofti*, *Brugia malayi* and *B. timori*) cannot be easily isolated from infected humans, since they reside in the lymphatics. The adult stage causes chronic diseases such as lymphedema and elephantiasis or hydrocele in men. Adult worms produce microfilariae (mf) that circulate in the blood and are readily available for genomic characterization. However, the amount of DNA obtained from a single mf is insufficient for genome sequencing. Such technical obstacles have restricted population-scale genomic analyses of field isolates in humans [6]. Earlier studies also depended on laboratory (animal) passages to obtain developmental stages (adults or L3s) that yield higher quantities of DNA [7, 8]. However, these methods can lead to a loss of genetic diversity and unintended changes in allele frequencies due to selection or genetic drift.

Genomic analysis of individual mf using Next Generation Sequencing (NGS) can, however, be achieved through DNA amplification. A 2019 study applied selective whole-genome amplification (SWGA) to *W. bancrofti* [6]. The SWGA approach utilizes isothermal amplification based on the highly processive Phi29 DNA polymerase, similar to the conventional whole-genome amplification (WGA) method. However, SWGA differs from WGA in that it uses primers based on nucleotide sequence motifs that are prevalent in the target genome (parasite) but infrequent in the background genome (host), rather than using random hexamers. This strategy selectively amplifies the target genome from samples in which it originally represented a minor fraction of the total DNA. While SWGA has been employed in genomic studies of bacteria and *Plasmodium* species from clinical samples [9], it is uncertain whether the method is efficient and reliable for multicellular eukaryotic pathogens such as filarial nematodes, which possess relatively large and complex genomes.

Similar to other helminth infections where the adult-stage parasites cannot be easily obtained from human hosts, the infection burden of filarial parasites is measured indirectly by counting larvae or through serologic means. These methods do not offer sufficient data to accurately estimate the number of reproductively active adult parasites or to quantitatively track changes in the adult worm population within the host. As a result, distinguishing between recrudescence (poor drug response leading to persistent infection) and new infections (ongoing transmission) after treatment using these indirect methods is challenging. Nonetheless, understanding the cause of rebound infections is crucial for permanent elimination. Within an area that passed the WHO recommended transmission assessment surveys and stopped MDA, it is possible that transmission may resume without the introduction of new infections [10, 11]. Reasons for this could include: (i) hidden active infections because of non-compliance to MDA, (ii) spontaneous resumption of mf production because treatment does not kill all adult worms, which have a long life expectancy, or (iii) reinfection from animal reservoirs (in the case of *B. malayi*). Resumption of effective MDA with the same or a more efficacious drug combination may solve the problem in these cases [12]. In contrast, transmission may also resume because of the introduction of parasites from other regions by infected mosquitoes that may fly or drift long distances, or because of immigrants from LF endemic areas [13]. For these cases, resumption of MDA in the entire implementation unit may not be needed if reintroduction of parasites can be prevented.

Genetic analysis of offspring parasites allows for the inference of adult parasite populations that are not directly accessible for examination [14]. Through kinship analysis, one can reconstruct family structures and estimate the number of breeding adults. By monitoring these families over time, insights into adult worm survival and fecundity can be gained. However, interpreting offspring genetic data requires careful analysis due to the complexity of parasite biology. For instance, the analysis can be influenced by polyandry [7, 15], density-dependent fecundity [16], and sampling biases resulting from reproductive skew among the breeders [17].

Here, we demonstrate that by analyzing the genetic variation in mf before and after drug treatment, we can infer the survival/fecundity of adult worms. This analysis enables the identification of parasites (and their genotypes) that respond differently to treatment. Such identification is vital when investigating the role of parasite genetics in treatment outcomes, such as suboptimal drug responses. This advancement marks a significant and crucial step towards genomic epidemiology and surveillance, which will be invaluable in controlling filariasis. Building upon this, we have developed *W. bancrofti* exon capture probes to facilitate the creation of field-deployable genotyping tools for estimating worm burdens and monitoring parasite population dynamics during MDA.

## Methods

### Parasite material – *Brugia malayi*

Adult *B. malayi* of the TRS strain [18] were produced in gerbils as described previously [19]. To generate DNA for experiments comparing different whole genome amplification (WGA) methods, adult males were collected and their DNA was isolated using the whole worm. Sixteen individual females were collected from 2 gerbils, washed in PBS, and cultured overnight in RPMI medium at 37°C. Thereafter, the apical end of the female (proximal to the genital opening) was removed and stored for isolation of maternal somatic DNA while minimizing DNA contamination from embryonic material. Microfilariae, which had been released into the RPMI medium, were collected from each female and stored at -20°C. After whole-genome sequencing analysis of the *B. malayi* females, 2 female worms displaying high levels of heterozygosity were selected, and 40 of their F_1_ progeny mf that had been collected as described above were selected for DNA isolation.

### Parasite material – *Wuchereria bancrofti*

Blood samples of *W. bancrofti* mf-positive subjects were collected before and after treatment during a clinical trial in southeastern Côte d’Ivoire, as previously described (Additional file 1: **Table S1 and S2**) [20, 21]. De-identified blood samples were shipped to Washington University School of Medicine in St. Louis and stored at -80°C. For isolation of individual mf, samples were thawed on ice. Depending on the mf concentration in the blood, 20-100 µl of blood was mixed with 900 µl PBS. Then, 200 µl of this solution was pipetted onto a slide and single microfilariae were picked under a Primostar dissecting microscope (Zeiss, Jena, Germany) at 4-fold magnification. The mf were collected in a 1.5 ml tube containing 200 µl fresh PBS. These mf were then placed onto a slide and single mf were picked and collected in a 1.5 ml tube containing fresh PBS. These washing steps were repeated 3-5 times, depending on the initial volume of blood. In the final step, single mf were picked from the slide using a micropipettor (10 µl volume), transferred individually to 0.2 ml microfuge tubes, and were either used for DNA extraction or stored at -20°C until further use.

### DNA isolation, whole genome amplification and sequencing

The DNeasy Blood and Tissue kit (Qiagen) was used for DNA isolation from adult *B. malayi* worms, followed by ethanol precipitation. For DNA isolation from single mf, we used the CGP protocol [22] with modifications: 25 µl of lysis buffer (950 µl nuclease-free water, 30 µl 3 M Tris-HCL, 5 µl NP-40, 5 µl Tween-20, and 10 µl proteinase K) was added to each tube containing one mf. The samples were then incubated at 55°C for minimum of 2 hours, followed by a 20 min 85°C proteinase K inactivation, and finally stored at 4°C. Successful DNA extraction was confirmed by qPCR using HhaI and LDR primers for *B. malayi* and *W. bancrofti*, respectively (Additional file 1: **Table S3**) [23, 24]. Positive samples were then amplified using the Ready-To-Go GenomiPhi V3 DNA Amplification kit (cytiva, Marlborough, MA) according to the manufacturer’s recommendations. After WGA, the sample was diluted 1:10, and the presence of parasite DNA was confirmed. For the quantification of human DNA, we used primers that amplify a section of Chromosome I [7].

Selective whole-genome amplification (SWGA) was carried out as previously described [25] with slight modifications. Primers were designed using the SWGA primer design toolkit, following the guidelines provided by the authors (Additional file 1**: Table S4**) [25]. Reactions were performed in a volume of 50 µl using input DNA (<1 ng/µl), 2.5 µM total of SWGA primers, 1× phi29 buffer (New England Biolabs, Ipswich, MA), 1× bovine serum albumin (BSA), 1 mM dNTPs, and 30 units phi29 polymerase (New England Biolabs). The amplification conditions were 35°C for 5 min, 34°C for 10 min, 33°C for 15 min, 32°C for 20 min, 31°C for 30 min, 30°C for 16 hours, followed by deactivation of the enzyme at 65°C for 15 minutes. Successful amplification was confirmed by qPCR, as was performed for the WGA samples. Amplified samples were directly used for library construction. Kapa Hyper PCR-free library was generated from the DNA sample and sequenced on Illumina’s NovaSeq platform (2 × 150bp paired-end reads) to ∼10Gb per sample.

### Analytical processing of the reads, variant calling, and kinship analysis

Sequencing reads were adapter/quality trimmed using trimmomatic v0.39 and were aligned against the combined reference assembly of the filarial worm nuclear, mitochondria, and the *Wolbachia* endosymbiont genomes using BWA v0.7.17 [26]. For clinical mf samples, the human genome was also included for sample quality assessment. Duplicate reads were removed and single-nucleotide variants (SNPs) were called using GATK v4.2.2 [27]. The following set of quality filters were applied to obtain high-confidence genotype calls in GATK: QD < 2; QUAL < 30; SOR > 3; FS > 60; MQ < 40; MQRankSum < -12.5; ReadPosRankSum < -8; DP > (2 × median depth) [28]. Variants were annotated according to their genomic locations and predicted coding effects using SnpEff v5.0c [29]. PLINK v1.9 [30] was used for performing principal component analysis (PCA) or multidimensional scaling (MDS) to determine sample relationship, and computing inbreeding coefficients (*F*_IS_) to identify samples with excessive heterozygosity (likely indicative of sample contamination). Kinship analysis was performed to identify familial relationships among the samples using both autosomal and X-chromosomal PCA. This analysis was guided by the kinship coefficients computed using the KING method [31], implemented in AKT v0.3.3 [32], and by the maximum-likelihood phylogenetic trees of the X-chromosome haplotypes constructed using IQ-TREE v2.2.0 (with the best-fit model automatically selected by ModelFinder) [33]. The local PCA approach, implemented in the *lostruct* package in R [34] (with a sliding window of 20 SNPs and a sum of squared values of the covariance matrix >50) was used to examine the X-chromosome haplotype structure. ChromoMap [35] was employed for haplotype visualization. The mitochondrial haplotype network was constructed in PopART [36] using the Minimum Spanning Network method [37].

### *Wuchereria bancrofti* exome sequencing and performance evaluation

*Wuchereria bancrofti* protein-coding gene annotation: a previously published *W. bancrofti* genome assembly (GenBank Accession: GCA_005281725.1) [6] was re-annotated using both *ab initio* and homology-based annotation pipelines. Repetitive elements were softmasked with RepeatMasker v4.0.9 using a species-specific repeat library created by RepeatModeler v1.0.11 [38]. *Ab initio* gene predictions were generated using BRAKER2 v2.1.0 [39], which was trained using *B. malayi* protein sequences (PRJNA10729). MAKER v2.31.10 [40] was subsequently run to refine these gene models and reduce false-positive gene predictions. *W. bancrofti* microfilaria cDNA library (GenBank Accession: SAMN00155125), RNA-seq transcripts [6] assembled using StringTie v1.3.6 [41], *B. malayi* (PRJNA10729) CDS sequences, and protein sequences from WormBase ParaSite Version 13 [42] (*Acanthocheilonema viteae* PRJEB1697, *B. malayi* PRJNA10729, *Dirofilaria immitis* PRJEB1797, *Litomosoides sigmodontis* PRJEB3075, *Loa loa* PRJNA246086, *Onchocerca volvulus* PRJEB513) were provided to MAKER as transcript/protein evidence. Gene predictions without supporting evidence were excluded with the exception of those encoding Pfam domains, as detected by InterProScan v5.19 [43], to balance sensitivity and specificity [40, 44]. Homology-based gene predictions were generated in GeMoMa v1.9 [45] using *B. malayi* gene models (PRJNA10729), and were combined with the BRAKER/MAKER *ab initio* gene predictions. For gene loci where both homology-based and *ab initio* gene predictions were available, homology-based gene models were selected in the final gene build. The completeness of the resulting gene set was assessed using BUSCO v5.4.7 with Nematoda specific single-copy orthologs (OrthoDB v10) [46].

### Exome capture panel, sequencing and evaluation

We designed custom hybridization capture reagents (Twist Bioscience, South San Francisco, CA) targeting the exonic regions of the *W. bancrofti* genome. A panel of 120bp tiling-probes was designed to cover CDS targets defined by the BRAKER/MAKER *ab initio* gene predictions (described above). Problematic repeat regions were empirically identified from *W. bancrofti* whole-genome sequencing data based on read mappability (>50% mapped bases with mapping quality <10). Probes that overlapped with these regions, as well as those that exhibited high sequence similarity to the human genome (hg38) were removed from the design, resulting in a set of 150,846 probes (Twist Bioscience Custom Design ID: TE-92630402). This probe set covered 89% of the 14.9 Mb protein-coding space of the updated gene build. Hybridization reactions (8-plex), Streptavidin beads binding and purification were performed following manufacturer’s instructions. Post-capture library pools were sequenced on Illumina’s NovaSeq platform (2 × 150bp paired-end reads). Sequencing reads were processed and variants were called as described above for the whole-genome data, but the maximum DP filtering (GATK) was not applied. BEDTools v2.30 [47] and Picard v2.26.2 (CollectHsMetrics and GenotypeConcordance) [27] were used to compute coverage and variant call metrics.

## Results

The ultimate goals of the study were to 1) infer the number of reproductively active adult worms in *W. bancrofti* infected subjects based on the genome-wide genetic diversity among individual mf within a host, and 2) differentiate between new and persistent *W. bancrofti* infections by genetically tracking the survival and fecundity of unobserved adults throughout the treatment process. We undertook a step-wise approach to achieve these two goals. We took advantage of our *B. malayi* – gerbil infection model (**Fig. 1A**) to inform our *W. bancrofti* methodological and analytical approaches (**Fig. 1B**). A schematic representation of the overall study design, which summarizes the source and purpose of each sample set, is presented in **Fig. 1**. Initially, we evaluated WGA and SWGA methods to amplify filarial worm genomic DNA for sequencing-based genotyping. *B. malayi* adult male samples were used to (i) obtain sufficient DNA so that sequencing libraries could be generated from both amplified DNA and unamplified source DNA for direct comparison and (ii) avoid non-diploid genotypes from embryos, which would be present if gravid female samples were used. After establishing confidence in our methods, we proceeded to analyze a population of clinical field isolates of *W. bancrofti* collected from a recent open-label randomized controlled drug trial in Côte d’Ivoire (CDI) [21]. Furthermore, by leveraging the *B. malayi* – gerbil model, we produced mf with a known pedigree. This guided us to develop and validate our maternal sibship identification method, which utilizes X-chromosome genetic diversity. By longitudinally sampling post-treatment mf, we assessed the temporal shifts in parasite infra-populations a year after administering a single dose of a triple-drug treatment consisting of ivermectin, diethylcarbamazine, and albendazole (IDA) [21]. Lastly, we developed a whole-exome capture panel for *W. bancrofti*, designed to enrich parasite nuclear DNA from lower-quality samples that may contain host DNA contamination.

**Fig. 1.**
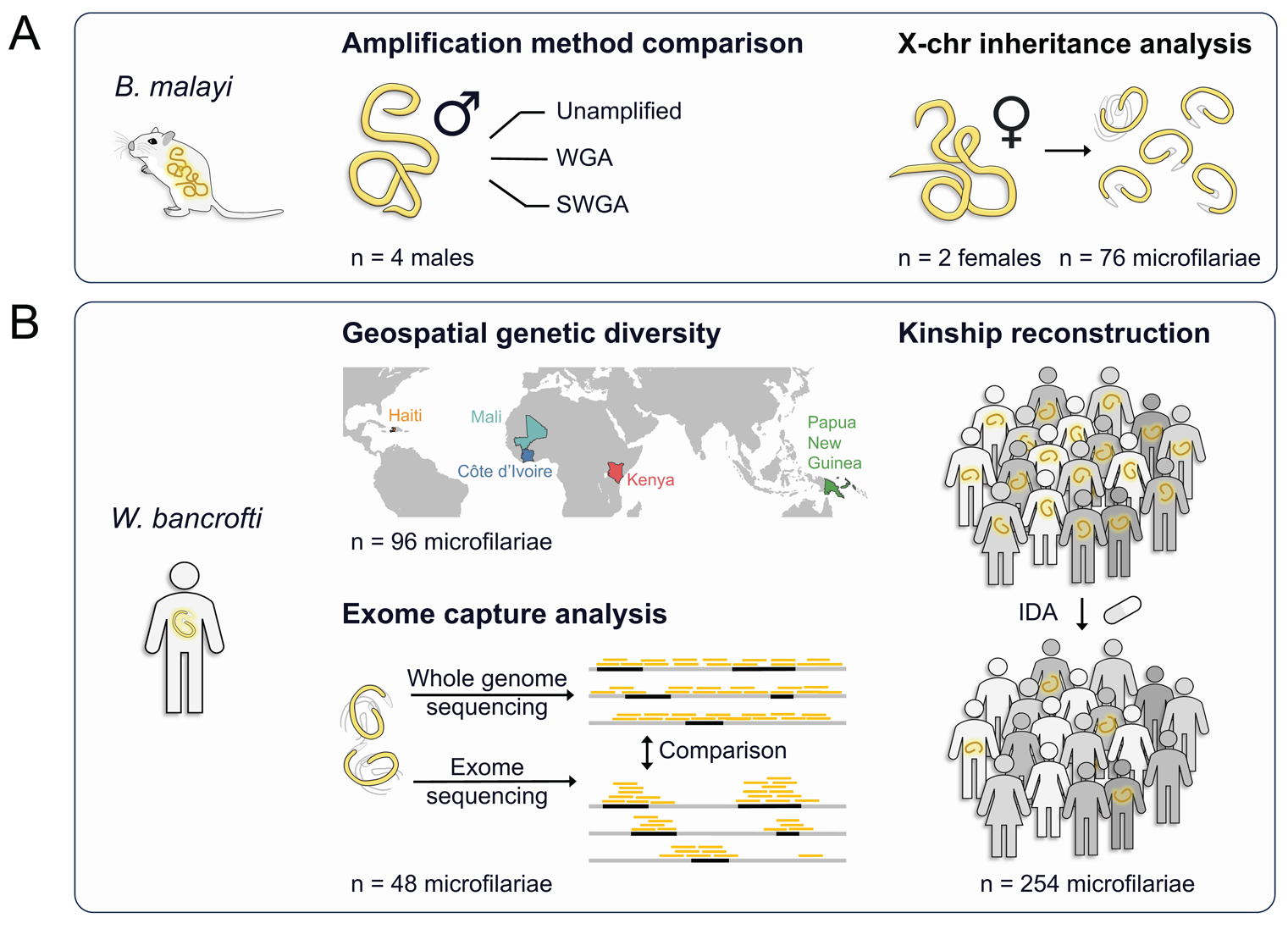
Overview of experiments and analysis. **A** *Brugia malayi* experiments. Adult worms collected from an experimental host (gerbils) and microfilariae of a known pedigree were used for methodology development. **B** *Wuchereria bancrofti* clinical isolates. Our genomics-based approach for analyzing worm burden was applied to samples obtained from human infections in Côte d’Ivoire, which were also utilized to develop probes for exome sequencing.

### Comparative analysis of genome coverage and genotyping reliability using different genome amplification methods for filarial DNA

Using DNA samples isolated from *B. malayi* adult males, we assessed the genome coverage bias and genotyping accuracy of WGA and SWGA methods relative to the unamplified DNA. SWGA primers were designed to minimize the distance between binding sites in the parasite genome and maximize the distance between binding sites in the host genome [25]. Three SWGA primer sets were tested on single-worm samples (SWGA1: manually curated set to improve primer sequence diversity; SWGA2: algorithmically-generated set prioritized based on the biding site distance; SWGA3: algorithmically-generated set prioritized based on the biding site evenness) (Additional file 1: **Table S4**). For the WGA method, random primers were used. We estimated the change in genome coverage with varying sequencing effort by down-sampling each library (**Fig. 2A**). For a given library size (i.e., total bases sequenced), DNA samples without amplification achieved better genome coverage than amplified samples, and WGA was more effective in amplifying *B. malayi* genomic DNA compared to SWGA. To achieve 90% coverage of the autosomes using WGA, sequencing of 15 million reads were required. To achieve a similar level of genome coverage using SWGA, 4 times more sequencing data (>60 million reads) were necessary. This implies that for samples containing up to ∼75% host DNA contamination, WGA would outperform SWGA (assuming that host and parasite DNA is amplified with equal efficiency in WGA and only parasite DNA is amplified in SWGA). Hemizygous sequences, such as the X chromosome in males, required ∼2 times more sequencing data compared to the diploid autosomes to achieve the same coverage. The mitochondrial genome was amplified effectively by both WGA and SWAG with minimal sequencing effort.

**Fig. 2.**
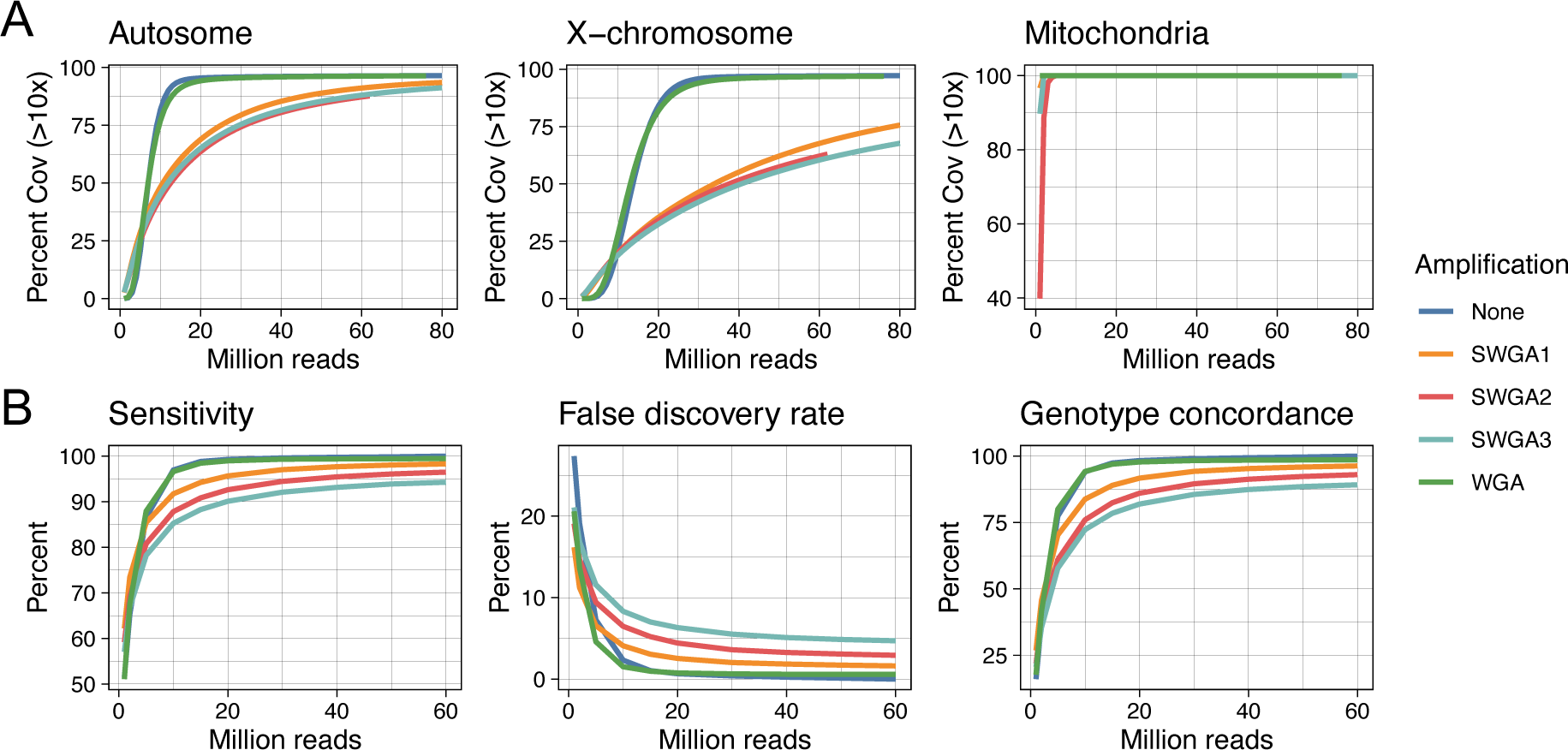
Performance assessment of genome amplification methods. **A** Sequencing effort needed to achieve >10x coverage across the target genome. **B** Evaluation of variant call sets using the unamplified sample (60 million reads) as the truth set.

We quantified genotyping errors as a function of amplification method and library size (i.e., overall sequencing effort) (**Fig. 2B**) to better understand the level of technical artifacts from WGA/SWGA with respect to the biological genetic variability (**Fig. 3**). Using the unamplified DNA sample as the ground truth, we evaluated the sensitivity, false discovery rate, and genotype concordance of the variant call sets generated by both the WGA and the SWGA method. The decrease in genome coverage associated with amplification (**Fig. 2A**) resulted in a lower sensitivity for SNP detection (**Fig. 2B**). In addition, the number of false positive variants and discordant genotypes increased when DNA samples were amplified using WGA or SWGA. Our data also showed that despite the increase in genotyping errors, samples clustered in PCA by worm and not by amplification method or library size (**Fig. 3**), indicating that genome-wide SNP data generated using amplified DNA correctly captured the genetic relationship between individual worms. This in turn suggested that temporal changes in (infra)population structure associated with treatment could be studied using WGA or SWGA approach.

**Fig. 3.**
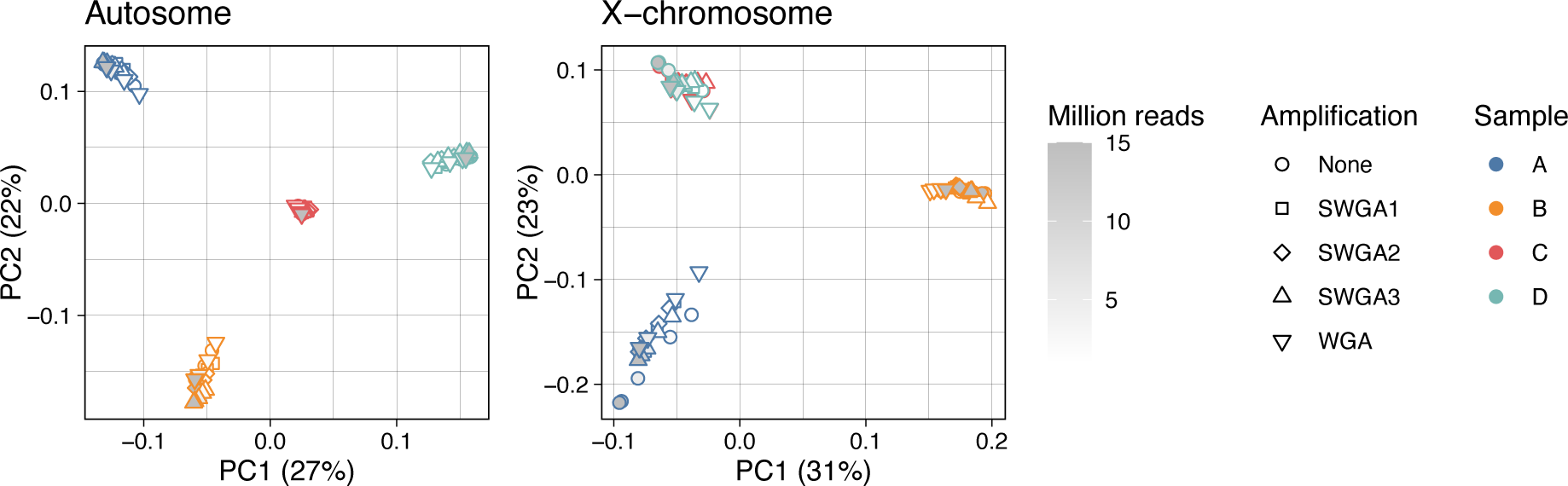
Principal component analysis of four individual *B. malayi* adult male worms. Genomic DNA samples were sequenced either without amplification or with WGA/SWGA. For each amplification group, variants were called after simulating 5 different library sizes (1, 2, 5, 10, 15 million reads). Each variant call set representing the unique combination of variables (worm, amplification method, and library size) were plotted as separate data points, and colored according to the worm (samples A-D). Adult male samples C and D had identical X-linked genotypes, suggesting that they are likely maternal siblings.

### Estimating the number of reproductively active adult parasites based on the genetic diversity of offspring microfilariae

We sequenced a total of 343 *W. bancrofti* mf samples collected from the IDA trial in Côte d’Ivoire (CDI) [21], including pre-treatment samples from 14 subjects (n=96) and pre- and 1-year post-treatment paired samples from 4 subjects (n=247). Guided by our *B. malayi* analysis which demonstrated that WGA achieved better genome coverage compared to SWGA, we used the WGA method to sequence individual *W. bancrofti* mf. Initial sequencing of the amplified DNA indicated that the level of host DNA contamination could be high and variable between samples (data not shown). Considering the 38-fold difference in genome size, even a small number of contaminating human cells can result in a significant number of human-derived reads in the final sequencing data. We therefore directed our efforts to improve the sample preparation method to minimize host cell contamination. Our approach (described in detail in the Methods section) included multiple washing steps and qPCR-based quality screening steps to ensure all samples have acceptable worm-to-human DNA ratios. On average, more than 95% of the autosome, 85% of the X-chromosome, and 99% of the mitochondrial genome were covered by at least 10x (**Fig 4A**).

**Fig. 4.**
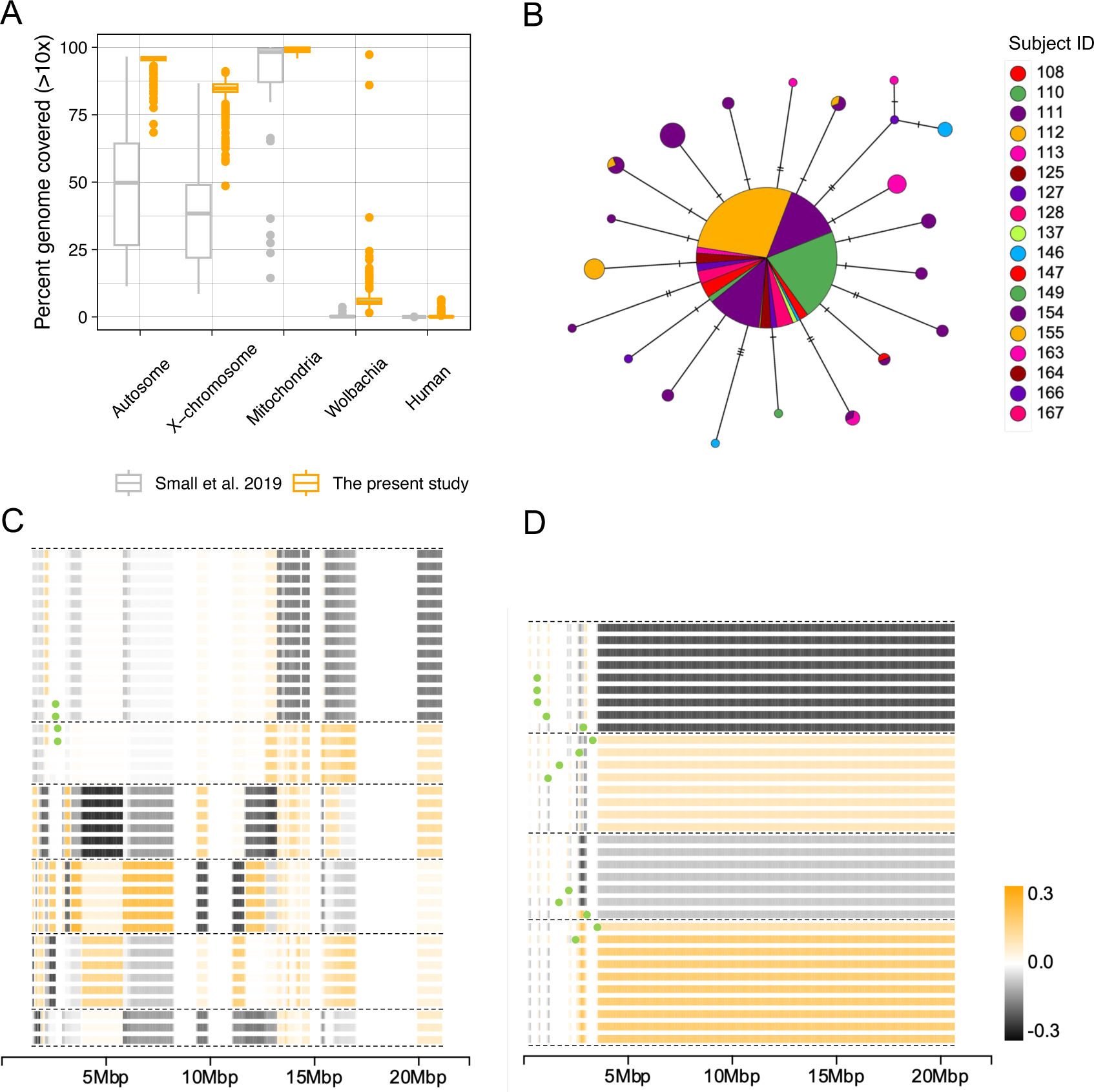
Analysis of mitochondrial and X-chromosomal genetic diversity using single microfilaria sequencing. **A** Improved genome coverage from single microfilaria sequencing using the optimized WGA protocol developed in this study, compared to previously published SWGA libraries by Small *et al.* [6]. **B** Mitochondrial haplotype network of 343 *W. bancrofti* mf collected from 18 subjects in Côte d’Ivoire. Male X-chromosome haplotypes inferred using “local” principal component analysis in **C** *W. bancrofti* mf collected from subject 155 (n=40), and **D** *B. malayi* mf collected from 2 adult females (n=34). Horizontal lines represent individual haplotypes. PCA was performed along the length of chromosome (excluding the pseudoautosomal region) in sliding windows of 20 SNPs. PC1 values were used to color each window. Recombination breakpoints were marked by green dots.

Because mitochondrial genomes (mtDNA) are maternally inherited, we first investigated whether we could estimate the number of adult female worms by analyzing the mitochondrial genetic diversity among mf sampled from each human subject. Sequence comparisons of the 13.5 kb mtDNA across all samples revealed low genetic diversity. We observed extensive haplotype sharing between unrelated mf collected from different subjects (**Fig. 4B**). These results indicate that mitochondrial data are not suitable for parentage inference. Like other filarial worms, *W. bancrofti* has an XY sex-determination system, and the inheritance pattern of the X-chromosome should allow the determination of maternal lineages using X-linked loci in male F_1_ progeny mf. Due to the hemizygous state of the male X-chromosome, haplotypes can be directly inferred from SNP data. By performing “local” principal component analysis (PCA) in sliding windows along the length of the chromosome, we could examine the long-range haplotype structure at the chromosomal level across a large number of mf samples (**Fig. 4C/D**). For instance, among the male mf collected from Subject 155 (n=40), we identified 6 major haplotypes, including 4 sequences representing recombinant haplotypes (**Fig. 4C**). Since a single diploid female worm can pass on 2 X-chromosome haplotypes (and their crossover recombinants) to the next generation, these results suggest that mf from Subject 155 descended from 3 female worms.

To better understand the inheritance patterns of X-chromosome genetic diversity in filarial parasites and validate our interpretation of the data, we generated genotype data from samples with known familial relationships using experimentally tractable *B. malayi* worms maintained in gerbils (TRS strain). Due to the low expected genetic diversity of this inbred laboratory strain, we sequenced somatic DNA samples from 7 adult females to select 2 female worms with higher levels of X-chromosome heterozygosity (Additional file 1: **Table S5**). Subsequently, the F_1_ progeny mf (n=76) of these females were genotyped. Using the sliding window PCA approach, we determined the X-chromosome haplotype structure among the male mf (n = 34). As expected, 4 major haplotype groups were identified, including their recombinant forms, consistent with 2 maternal lineages (**Fig. 4D**). These data also suggested that meiotic recombination predominantly occurred at the distal ends of the X-chromosome (proximal to the telomeres) in both *W. bancrofti* and *B. malayi*. Analysis of the haplotype structure, excluding these regions of high recombination rates, can provide complementary information helpful for determining the number of unique haplotype groups that are informative for maternal parentage inference.

### Discriminating persistent vs. new *W. bancrofti* infections based on kinship analysis of microfilariae before and after treatment

Next, we examined changes in the parasite infra-population, and investigated if the post-treatment populations likely represent a new or persistent infection (**Fig. 5**). Pairwise fixation index (*F_ST_*) analysis (Additional file 1; **Table S6**) indicated that there were varying degrees of genetic shift in parasite populations in each patient at 1-year post-treatment, ranging from a small change (e.g., subject 149) to a larger change (e.g., subject 111). We reconstructed familial relationships among individual microfilariae (**Fig. 6A**). We first determined X-chromosomal haplotype groups using the male mf (Additional file 2: **Fig. S1**), and then assigned these to maternal sibling families based on their autosomal genetic relationships (Additional file 1: **Table S7**), guided by a matrix of kinship coefficients (Additional file 1: **Table S8**). The analysis revealed persistent infections in subjects 149, 154, and 155, where individual mf from both pre- and post-treatment populations belonged to the same maternal family. In contrast, for subject 111, none of the post-treatment mf showed a familial relationship with any of the pre-treatment mf, indicating a new infection. Based on the maternal sibling groups, we estimated the minimum number of reproductively active adult females to range from 3 in subject 155 to 9 in subject 111 (**Fig. 6B**).

**Fig. 5.**
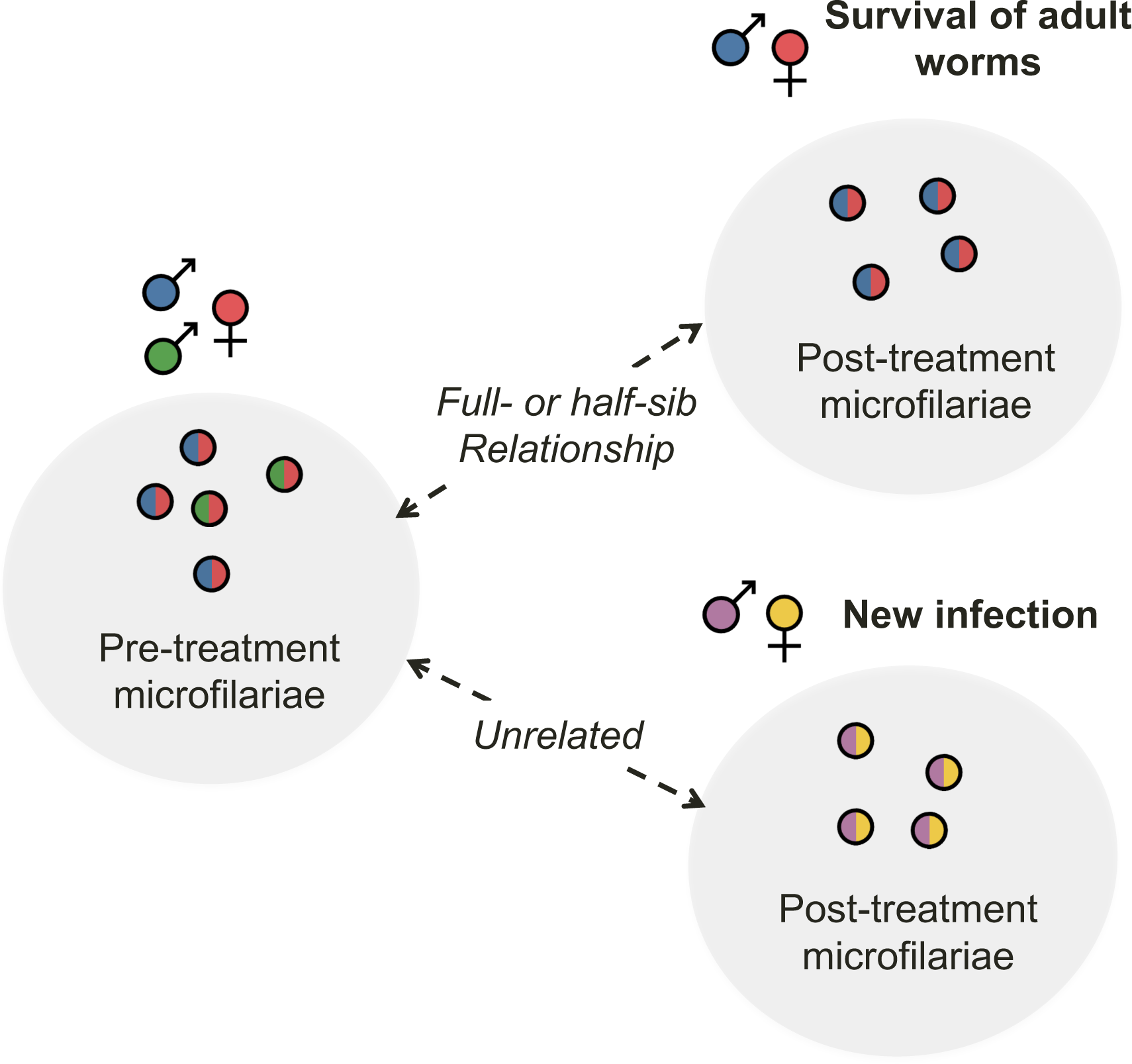
Schematics for identification of treatment failure and reinfection based on kinship analysis of offspring worms. The presence of familial relationships across pre- and post-treatment microfilariae indicates survival of adult worms and the absence of familial relationships indicates newly acquired infection.

**Fig. 6.**
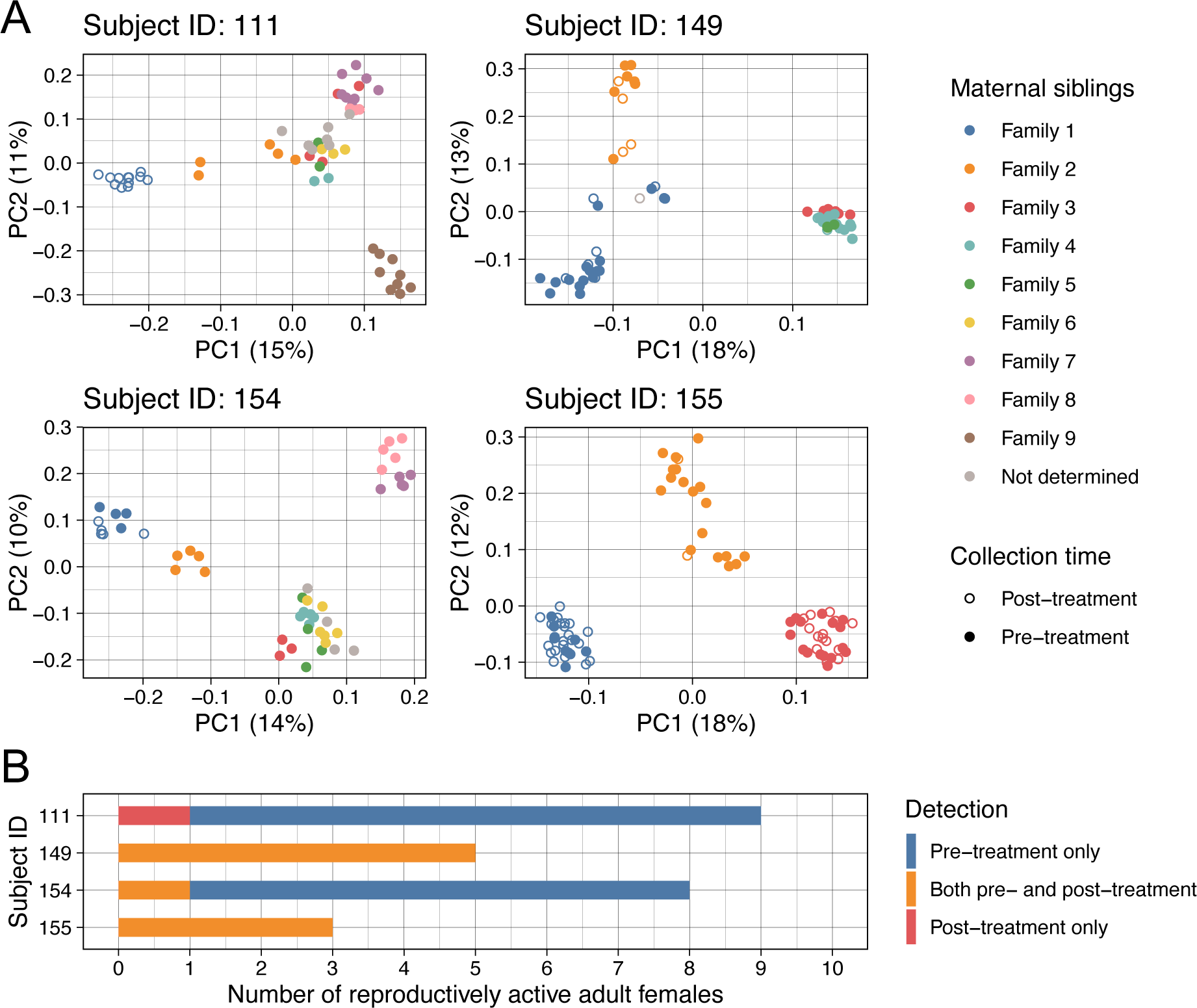
Monitoring worm burden based on the infra-population genetic diversity in *W. bancrofti*. **A** Maternal sibship relationship among *W. bancrofti* microfilariae. Kinship was inferred across pre- and post-treatment samples in each subject using autosomal, mitochondrial, X-, and Y-chromosomal SNPs. **B** Estimation of the minimum number of reproductively active female breeders based on the mf maternal sibling groups.

### *W. bancrofti* genetic diversity across geographic regions

We examined the population structure of *W. bancrofti* in our study site in Côte d’Ivoire (Agboville District) to investigate the possibility of identifying migrant parasites from ancestrally diverse populations. In addition to the 4 subjects from whom we collected paired samples both pre-treatment and 1-year post-treatment (n=254), we generated pre-treatment mf genomic data (n=96) collected from 14 additional subjects from CDI. To put the genetic diversity of these CDI samples into context, we conducted a principal component analysis incorporating previously published samples from Haiti, Kenya, Mali, and PNG (**Fig. 7A**) [6, 7]. The 96 pre-treatment mf from 14 subjects, along with 103 mf from subjects 111 and 154, formed a single cluster that likely represented the local parasite population in the Agboville District (**Fig. 7B**). Mf from subject 155 appeared genetically distinct from the rest of the Agboville population. In subject 149, mf belonging to maternal families 1 and 2 had genotypes characteristic of the Agboville population (PC2 > -0.05), while those belonging to maternal families 3, 4, and 5 had distinct genotypes (PC2 < -0.1). When CDI samples were compared to samples from other countries, they formed two distinct clusters: one representing the local Agboville population (PC2 < 0), and the other representing the mf from subject 155 (PC2 > 0.05) (**Fig. 7A**).

**Fig. 7.**
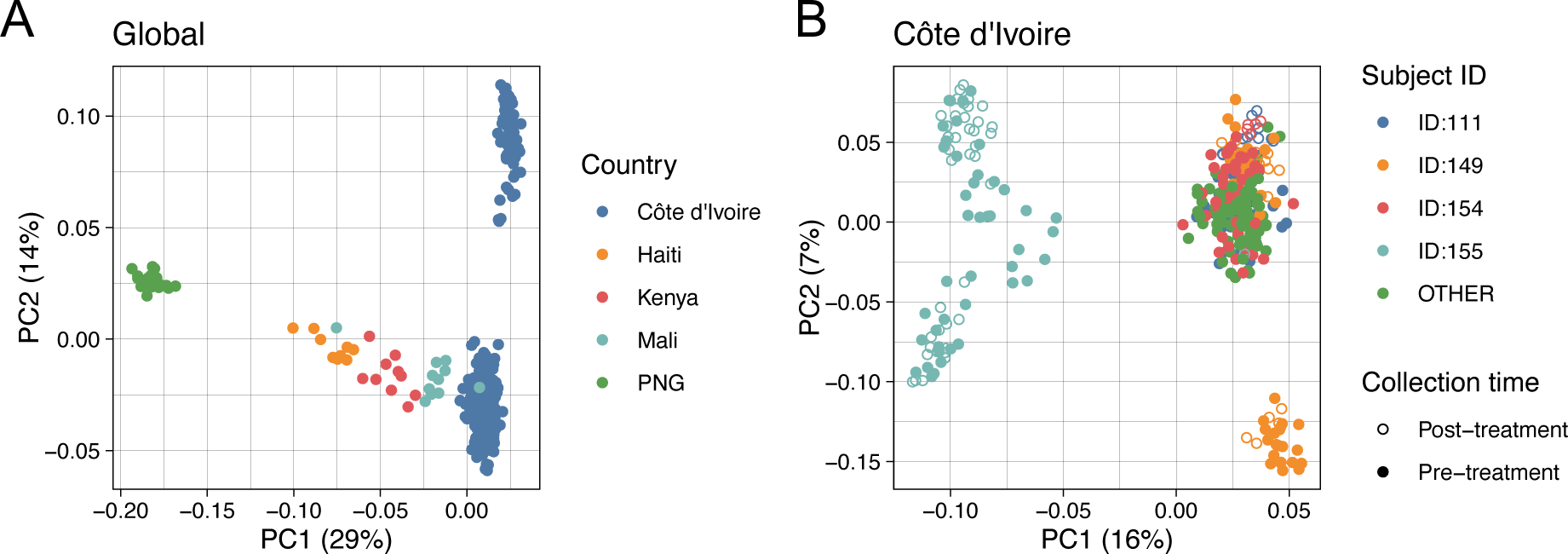
Principal component analysis (PCA) of *W. bancrofti* genetic diversity. All publicly available genomic data for *W. bancrofti* [6, 7] were included to contextualize the genetic diversity of the samples collected from Côte d’Ivoire.

### *W. bancrofti* whole-exome sequencing and performance evaluation

In preparation for developing a targeted sequence capture panel to facilitate sequencing of mf DNA in samples with high human contamination, we first improved the annotation of the existing genome assembly. The reference genome of *W. bancrofti* was updated by Small *et al.* [6] with a significant improvement in the contiguity over the previously published assembly. However, the quality of protein-coding gene annotation remained relatively poor. We re-annotated the genome to improve the accuracy of gene models. After re-annotation, the mean number of exons per gene increased from 5.3 to 9.1, and the mean CDS length increased from 0.73 kb to 1.3 kb (**Table 1**). The detection of full-length BUSCOs (highly-conserved core *Nematoda* genes) [48] increased from 47.5% to 98.4%, indicating a substantial number of incomplete, fragmented or missing gene models were corrected in our updated gene set (**Table 1**).

**Table 1.** Genome annotation and completeness statistics (based on longest isoforms) of *W. bancrofti*, related filarial species and the model nematode *C. elegans*.

|  | *Wuchereria bancrofti* |  | *Brugia malayi* | *Loa loa* | *Onchocerca volvulus* | *Caenorhabditis elegans* |
| --- | --- | --- | --- | --- | --- | --- |
|  | Small 2019 | Re-annotation |  |  |  |  |
| BioProject | PRJNA275548 |  | PRJNA10729 | PRJNA37757 | PRJEB513 | PRJNA13758 |
| Total genome length (bp) | 88,416,250 |  | 88,235,797 | 91,373,458 | 96,427,137 | 100,286,401 |
| Number of protein-coding genes | 9,651 | 11,166 | 10,878 | 14,908 | 12,109 | 19,981 |
| Total CDS length (bp) | 7,063,809 | 14,859,837 | 14,876,705 | 14,767,276 | 14,979,205 | 24,568,839 |
| Mean CDS length (bp) | 732 | 1,331 | 1,368 | 991 | 1,237 | 1,230 |
| % of genome covered by CDS | 8 | 16.8 | 16.9 | 16.2 | 15.5 | 24.5 |
| Mean exon length (bp) | 144 | 147 | 160 | 150 | 199 | 227 |
| Mean intron length (bp) | 358 | 371 | 355 | 333 | 431 | 327 |
| Mean exons per mRNA | 5.3 | 9.1 | 9.1 | 6.6 | 8.8 | 4.6 |
| Complete BUSCOs | 47.5% | 98.4% | 98.9% | 98.2% | 98.2% | 100.0% |
| Complete and single-copy | 46.9% | 96.8% | 98.3% | 97.9% | 97.2% | 99.6% |
| Complete and duplicated | 0.5% | 1.6% | 0.6% | 0.3% | 1.1% | 0.4% |
| Fragmented BUSCOs | 6.2% | 0.3% | 0.2% | 1.5% | 0.5% | 0.0% |
| Missing BUSCOs | 46.3% | 1.3% | 0.9% | 0.3% | 1.3% | 0.0% |

We designed custom hybridization capture probes (120 bp) comprised of 150,846 oligonucleotide baits targeting the coding exome of *W. bancrofti*. The design covered 13.2 Mb of 14.9 Mb total coding exome (88.7%) and 1.6 Mb of adjacent intronic/intergenic regions. Capture experiments were performed to evaluate the effectiveness of parasite nuclear genome (nDNA) enrichment using 48 single mf libraries containing a range of human DNA (up to 48.7%) and *W. bancrofti* mtDNA (up to 50.0%). On average across all samples, 95.1% of mapped bases were localized on or near (<250bp) the regions targeted by the probes (i.e., the baited regions), indicating an overall high on-target rate. A mean coverage of 427.5x was achieved across these baited regions, representing a 128-fold enrichment above the genomic background. We compared the sequence composition of WGS and exome capture libraries derived from the same single mf source DNA (**Fig. 8A**). The hybridization capture approach effectively enriched *W. bancrofti* nDNA sequences. In addition, we observed a >90-fold reduction of human derived reads after exome capture among the samples that contained >5% human derived reads (n=20). Similarly, an >18-fold reduction of mtDNA was observed among samples that initially contained >5% mtDNA (n=28).

**Fig. 8.**
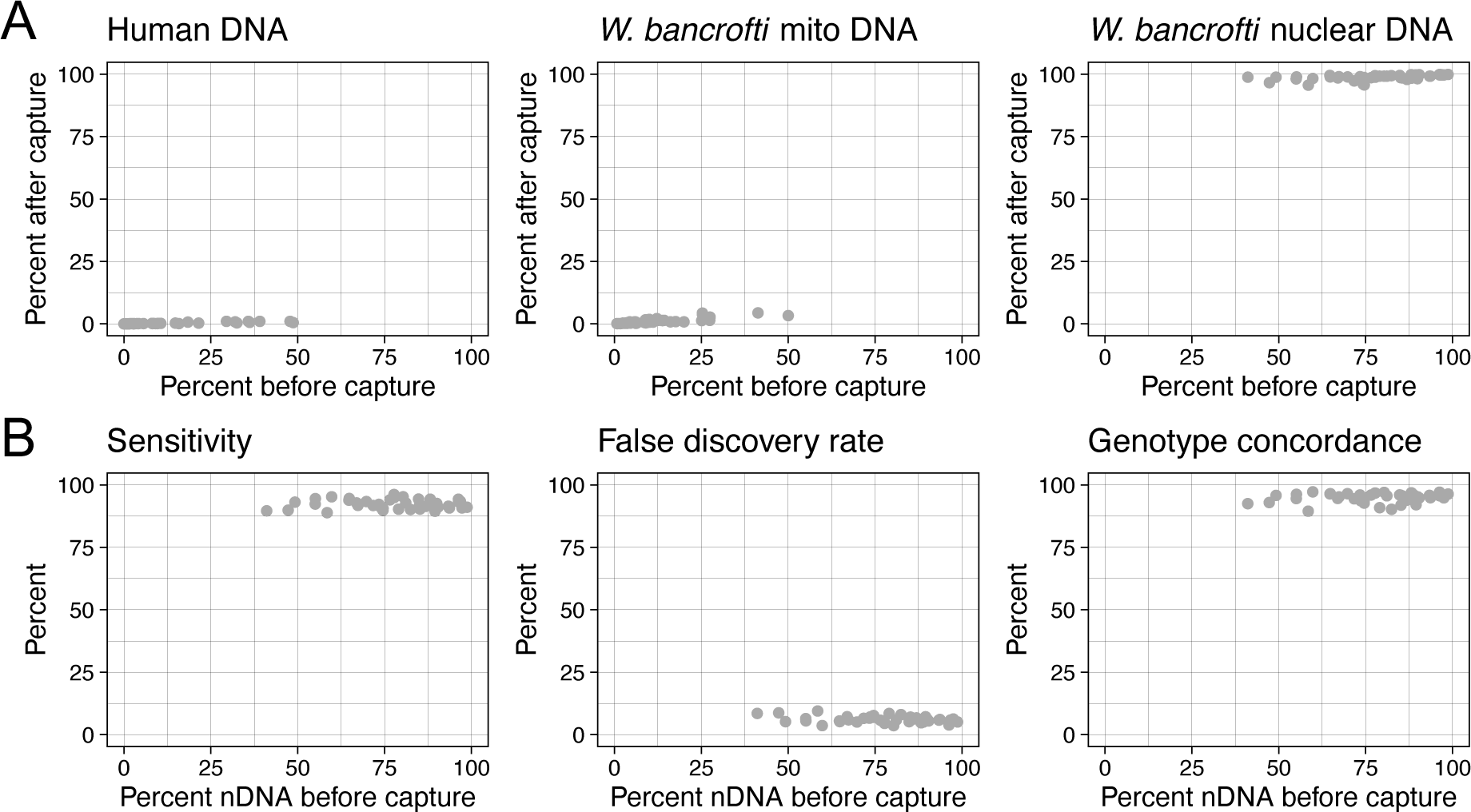
Performance assessment of *W. bancrofti* exome sequencing. **A** Reduction of human and mitochondrial DNA and enrichment of parasite nuclear DNA (nDNA) by hybridization capture of sequencing libraries. **B** Accuracy of variant calling from exome data.

We called SNP variants using the exome data and assessed the accuracy of the resulting genotypes relative to the call set generated from the whole-genome data that served as the truth-set. On average across the 48 samples, ∼9K non-reference variants were called within the baited coding regions per sample. The mean sensitivity, false discovery, and genotype concordance rates were 92.8%, 4.9% and 96.0%, respectively. We then investigated whether the pre-capture library composition significantly affected these genotype metrics (**Fig. 8B**). To minimize confounding due to variation in read coverage between samples, we subsampled the exome sequencing data to a uniform library size prior to variant calling. The mean sensitivity, false discovery, and genotype concordance rates of the variant call set from the subsampled exome libraries were 92.3%, 6.0% and 95.0%, respectively. Within the range tested, the genotyping quality of exome data was minimally affected by the amount of human and/or *W. bancrofti* mitochondrial DNA present in the sample (**Fig. 8B**).

## Discussion

This study marks the first successful effort to overcome technical challenges that have historically impeded population-scale genomic analysis of LF-causing worms. Our initial step was to identify a method that could efficiently amplify the filarial worm genome without compromising subsequent genetic analysis. We evaluated genome coverage patterns and found that whole-genome amplification (WGA) outperformed selective whole-genome amplification (SWGA). SWGA introduced greater coverage bias than WGA, requiring four times more sequencing data than WGA to achieve similar genome coverage. For this reason, we optimized our single mf sequencing method based on WGA. Although both WGA and SWGA have been used for filarial parasites in the past, a stringent comparison regarding their efficiency and accuracy had not been conducted [6, 49]. By quantifying genotyping accuracy in relation to the amplification method, we observed an increase in the number of false-positive variants and discordant genotypes when using either WGA or SWGA. Despite this, samples clustered in PCA based on the worm from which the DNA was isolated, rather than by the amplification method or library size. This indicates that genome-wide SNP data generated from amplified DNA can accurately capture the genetic relationships between individual worms. These comparative data will also be valuable in future studies for determining whether specific genetic analyses (such as genome-wide ROH analysis, PSMC modeling, etc.) are sensitive to genotyping errors introduced by WGA or SWGA.

Population genomic studies based on sequencing individual filarial worms are very limited, and the analyses have mainly been based on mtDNA or a very low breadth of coverage of the nDNA [50, 51]. To improve nDNA coverage, our approach incorporated multiple washing steps and employed qPCR-based quality screening to ensure all samples had acceptable worm-to-human DNA ratios. For the first time, our results report analyses that cover, on average, more than 95% of the autosome, 85% of the X-chromosome, and 99% of the mtDNA with at least 10x coverage. This represents a significant improvement over previously published datasets generated using SWGA [6] and allows for the first comprehensive genome-wide variant analysis of individual mf in *W. bancrofti*.

This improvement enabled us to test and demonstrate methods for estimating the number of reproductively active adult worms in infected subjects, based on the genome-wide genetic diversity among individual mf within a host. We used *B. malayi* mf of known pedigree as a guide for interpreting the *W. bancrofti* data. Using both the autosomal and X-linked SNPs, we conducted kinship analyses to reconstruct familial relationships among individual mf. These analyses provided a quantitative measure of infection burden, specifically the number of active female breeders, even when the mitochondrial genetic diversity was too low for identifying maternal lineages. Efforts to eliminate LF are guided by transmission modeling using several models [52–55]. However, these models are based on empirical estimates of reproductively active female worms. The ability to provide experimental data on the number of female worms both before and after treatment could greatly enhance the accuracy of these models.

Our approach enabled us to genetically track the survival and fecundity of unobserved adults throughout the treatment process, differentiating between new and persistent infections. Analysis of our CDI *W. bancrofti* mf data suggested reinfection in one subject and recrudescence in three others. The fact that we discovered a subject with new infection indicates ongoing transmission, despite the low prevalence of *W. bancrofti* in the area and ongoing MDA. Non-compliance with MDA is a major problem for the Global Program to Eliminate LF and should be specifically addressed in the CDI national program [56]. In parallel, we also detected treated subjects with persistent infections (SOR parasites). This is not surprising because in CDI, only about 71% of infected subjects are free of mf after 12 months of IDA triple-drug treatment [21]. It is important to note that efficacy of IDA shows geographical differences; for example, in Papua New Guinea (PNG), 96% of treated subjects are free of mf 12 months after treatment [57]. Our whole genome data show that mf from CDI can be clearly differentiated from mf from PNG and it should be theoretically possible to determine whether a less treatment-susceptible strain from West Africa has been introduced to PNG.

Analysis of offspring genetic data to characterize the (unobserved) adult parasite population can be complicated by polyandry [7, 15] and sampling biases stemming from reproductive skew among the breeders [17]. Additionally, the number of offspring parasites required for a reliable analysis depends on the genotype diversity and frequency within the infra-population, which can vary substantially between hosts and is sometimes difficult to predict in advance. Inadequate/under sampling can lead to erroneous inferences about the survival/fecundity of the adults. As with all methods that rely on offspring worms, our approach cannot detect infections during the prepatent period (interval between the entry of the L3 larvae and the appearance of detectable mf), which can last several months in filariasis. Therefore, temporal sampling should be carefully designed to meet the requirements of specific research goals.

The analyzes of infra-population genetic variation of parasites within our study site in Agboville also identified a few mf that were genetically distinct from the rest of the local population. These formed a separate cluster when principal component analysis (PCA) was performed, even when including outgroup parasites from Haiti, Kenya, Mali, and Papua New Guinea (PNG). These likely represent migrating parasites associated with the long-distance movement of human hosts. At present, it is not possible to identify the precise source population(s) of these parasites based on their genetic profiles. However, as the database of geographically diverse *W. bancrofti* samples expands in the future, it may become possible to identify the source population and spatially track parasite movement associated with host/vector migration. Furthermore, our analysis helped identify mf that are the progeny of SOR parasites (e.g., those in subjects 149 and 155), as well as progeny from good responders (e.g., pre-treatment mf in subject 111). This lays the groundwork for future genome-wide association analyses (once a sufficient sample size is reached) to understand the genetic factors that influence variations in parasite drug response and to identify genetic markers for surveillance.

Finally, we incorporated multiple layers of quality control steps into our WGA pipeline to exclude samples with high levels of host contamination and/or DNA degradation. Residual human DNA in samples can result in lower worm genome coverage, and small circular genomes (e.g., mitochondrial genome) can be preferentially amplified during WGA when the input DNA is fragmented. However, samples from field isolates are frequently suboptimal due to storage and/or shipping conditions. To facilitate the use of most clinical field samples, we developed a whole-exome capture panel that enriches for *W. bancrofti* nDNA from a mixed sample containing host DNA. While several *W. bancrofti* genome assemblies with increasing contiguity have been published [6, 7, 58], the quality of the gene models was insufficient for designing high-quality exon capture probes. Therefore, we re-annotated the *W. bancrofti* genome and improved the BUSCO completeness score to 98.4%. By systematically comparing library compositions and genotyping accuracies, we demonstrated that a hybridization capture approach can be employed to achieve unbiased sampling of mf without the need to exclude samples with high levels of human DNA contamination and/or those of suboptimal quality. In the future, we anticipate that targeted genotyping approaches such as hybridization capture or multiplex PCR-based methods can be designed to focus on a smaller set of “informative” SNPs for sibship inference and parentage analysis. These methods would offer improved throughput and cost-effectiveness. Developing a well-curated database of genetic variations in clinical samples will then be crucial for understanding changes in parasite populations following drug treatment and interventions. Such a database, encompassing both global and regional genetic variation, will support the identification of possible source populations in cases of reinfection. Additionally, given a large enough sample size, the database could facilitate a genome-wide association study (GWAS) to investigate the contribution of parasite genetics to treatment outcomes.

## Conclusions

Using a combination of laboratory-produced *B. malayi* samples and field-collected *W. bancrofti* samples, we showed that whole-genome amplification is more effective and accurate than selective whole-genome amplification for the genomic analysis of mf. We provide evidence that analyzing mitochondrial genomes is insufficient for differentiating persistent infections from newly acquired ones; however, this differentiation can be reliably achieved through the analysis of the nuclear genome. Targeted exon capture is a robust approach for enriching the coding sequences of filarial parasites from lower-quality DNA samples. Our study lays the groundwork for developing field-deployable genotyping tools to estimate worm burdens and monitor parasite populations, which will be invaluable for the successful implementation of mass drug administration programs aimed at eliminating lymphatic filariasis.

## Data Availability

The dataset supporting the conclusions of this article is available in the NCBI Sequence Read Archive (SRA, https://www.ncbi.nlm.nih.gov/sra) under the study accession number SRP463861. Sample metadata is available in Additional file 1: Table S9.

## Abbreviations

CDI: Cote d’Ivoire
GWAS: Genome wide association study
IDA: Ivermectin, diethylcarbamazine, and albendazole
L3: Third larval stage
LF: Lymphatic filartiasis
MDA: Mass drug administration
mf: Microfilaria
mtDNA: Mitochondrial genome
nDNA: Nuclear genome
NGS: Next generation sequencing
PCA: Principal component analysis
PNG: Papua New Guinea
PSMC: Pairwise sequentially Markovian coalescent
ROH: Runs of homozygosity
SNP: Single nucleotide polymorphism
SOR: Suboptimal response
SWGA: Selective whole genome amplification
WGA: Whole genome amplification

## Declarations

### Ethics approval and consent to participate

Ethical approval (IRB ID #: 201103313), full waiver of HIPAA Authorization (IRB ID #: 201103313), and non-human subjects determination (IRB ID #: 201712119) were awarded by the Washington University in St. Louis Institutional Review Board (DHHS Federalwide Assurance #FWA00002284). All animal experiments were carried out under protocols approved by Washington University School of Medicine (20-0503) Institutional Animal Care and Use Committees (IACUC). All housing and care of laboratory animals conformed to the National Institutes of Health (NIH) Guide for the Care and Use of Laboratory Animals in Research (see 18-F22). Euthanasia was accomplished by CO_2_ inhalation.

### Consent for publication

Not applicable.

## Availability of data and materials

The dataset supporting the conclusions of this article is available in the NCBI Sequence Read Archive (SRA, https://www.ncbi.nlm.nih.gov/sra) under the study accession number SRP463861. Sample metadata is available in Additional file 1: **Table S9**.

## Acknowledgments

We thank the field team in Côte d’Ivoire for collecting blood samples from patients naturally infected with *W. bancrofti,* Yuefang Huang for providing laboratory produced samples of *B. malayi,* Devyn Yates for laboratory assistance and the McDonnell Genome Institute for genome sequencing.

## Authors’ contributions

Conceived and designed the study: PUF and MM

Collected specimens: MA and BGK

Biological and sequence data database maintenance: YC

Sample preparation, extraction of DNA, WGA and SWGA and qPCR: KF

Performed data analysis: YC

Interpreted the data: YC, PUF and MM

Wrote the paper: YC, PUF, KF and MM

All authors approve the manuscript for publication.

## Supplementary Information

### Additional file 1

**Table S1.** Patient information. **Table S2**. Parasite material. **Table S3**. Primers used for qPCR. **Table S4.** Primer sequences used for SWGA. **Table S5.** X-chromosome heterozygosity in *B. malayi* adult females (TRS strain). **Table S6.** Fixation index (*F*_ST_) analysis comparing pre- and post-treatment *W. bancrofti* mf populations. **Table S7.** Principle component analysis and kinship inference of pre- and post-treatment *W. bancrofti* mf populations. **Table S8**. Pairwise Kinship coefficients. **Table S9.** Sample metadata.

### Additional file 2

**Fig. S1.** Maximum-likelihood phylogenetic trees of the X-chromosome haplotypes in *W. bancrofti* male microfilariae.

