## Additional file 2 for "Distinguishing recrudescence from reinfection in lymphatic filariasis: a genomics-based approach for monitoring worm burden"

**Fig. S1.** Maximum-likelihood phylogenetic trees of the X-chromosome haplotypes in *W. bancrofti* male microfilariae.

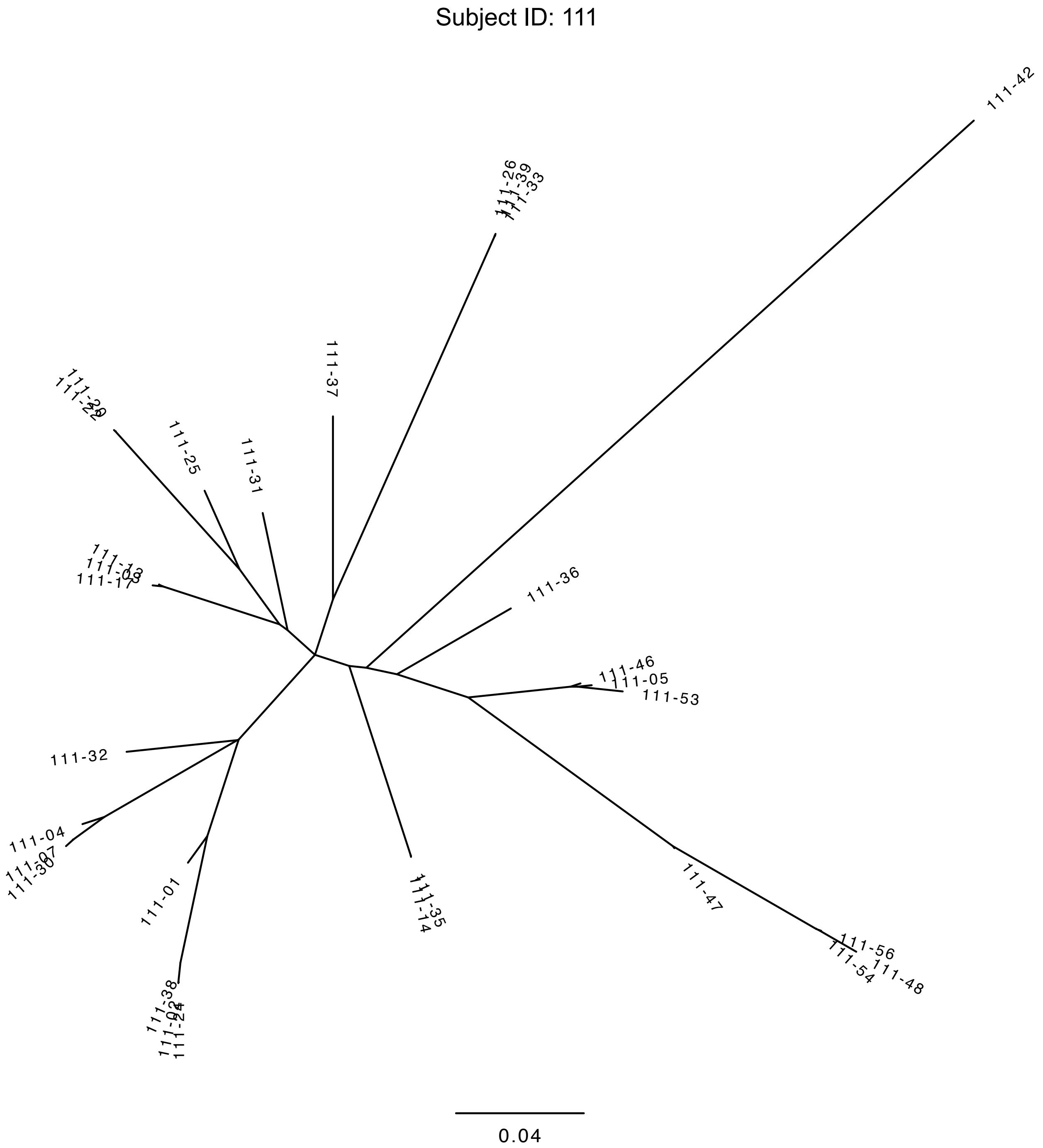

Subject ID: 149

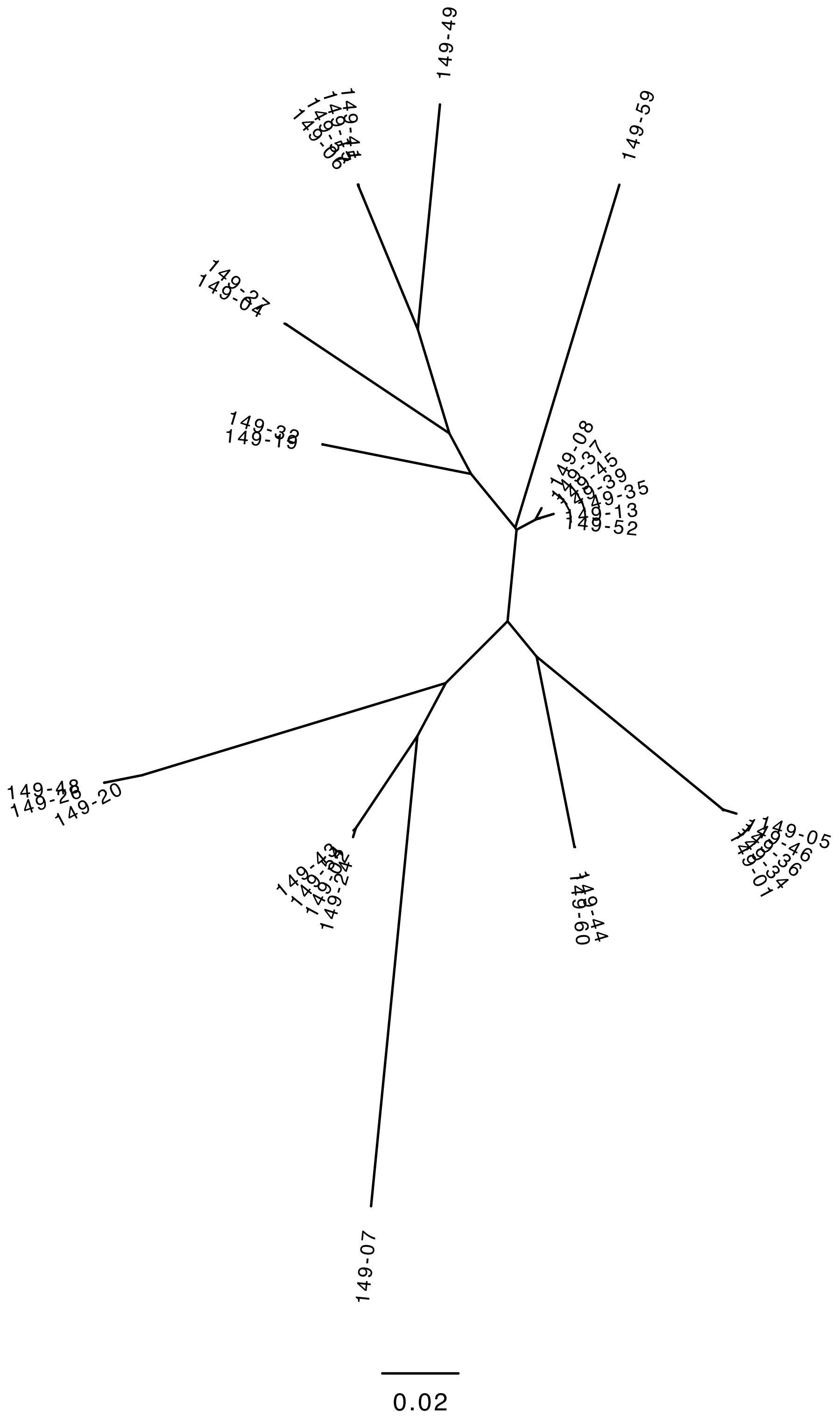

Subject ID: 154

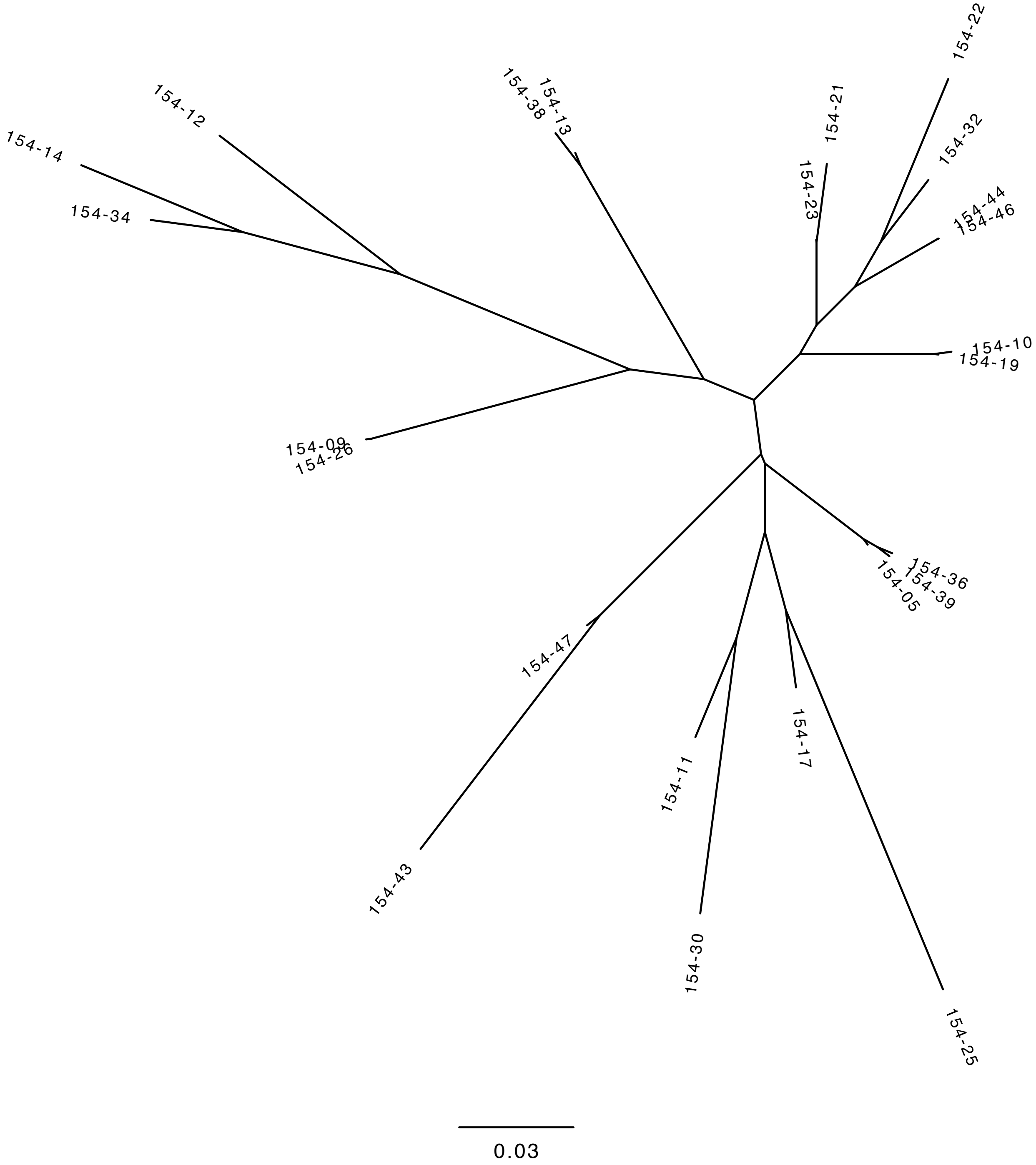

Subject ID: 155

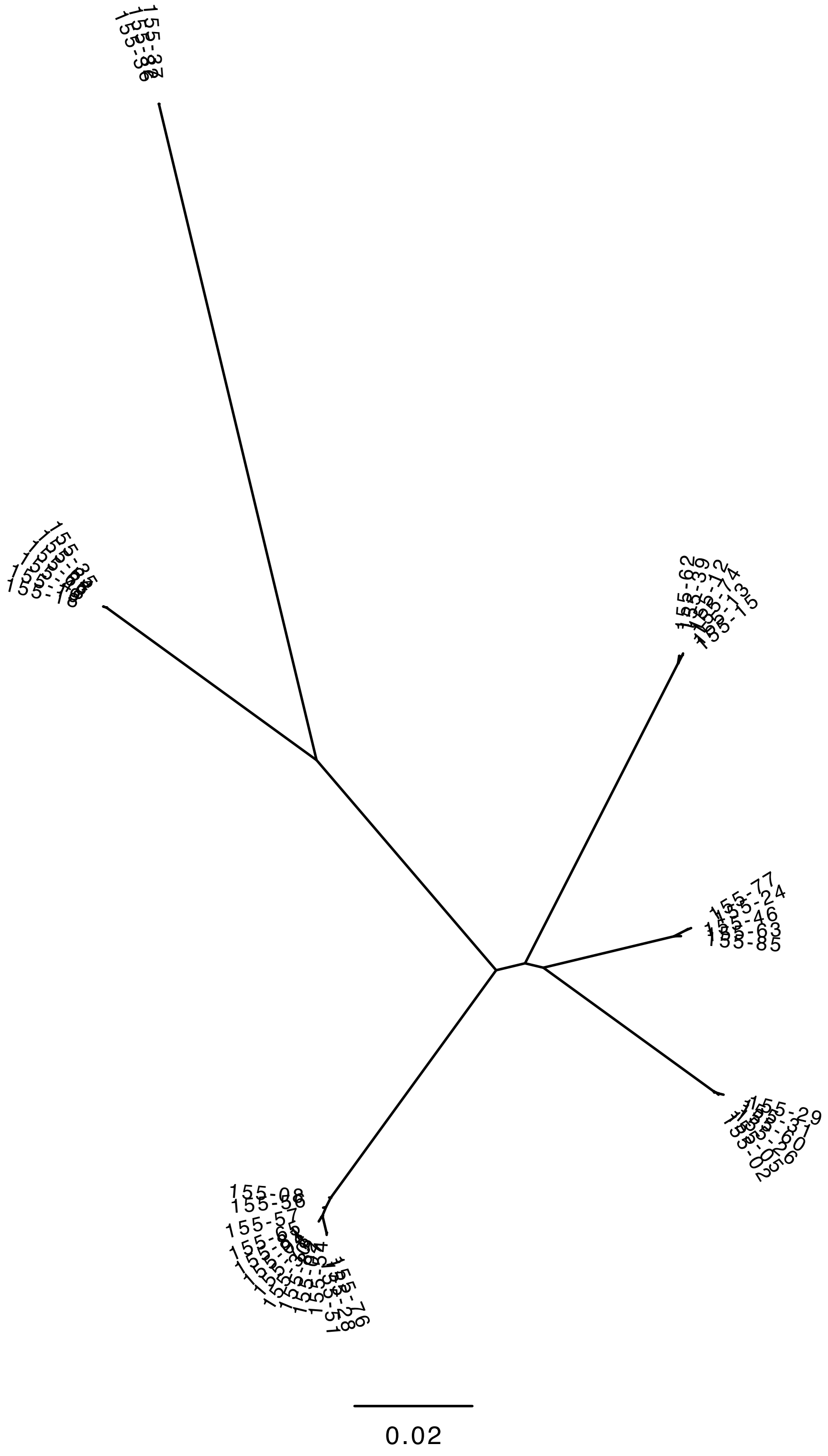
